# Impact of long-term care facility size on preparedness and adherence to infection prevention and control guidance for the mitigation of COVID-19

**DOI:** 10.1101/2021.06.12.21258774

**Authors:** Adherence to COVID-19 guidance, Patrick Alexander Wachholz, Ruth Caldeira de Melo, Alessandro Ferrari Jacinto, Paulo Jose Fortes Villas Boas

**Author notes:** <u>Corresponding author</u>: Botucatu Medical School, Universidade Estadual Paulista (UNESP), Av. Prof. Mário Rubens Guimarães Montenegro, s/n - UNESP - CEP 18618-687, Botucatu, São Paulo, SP, Brazil; Pabx: (14) 3880-1001, Twitter handle: @DrPatrickLTC. <u>Authors’ contributions</u>: Study conception and design: PAW, RCM, PJFVB; Acquisition of data: PAW, RCM, AFJ, PJFVB; Analysis and interpretation of data: PAW, RCM; Drafting of the manuscript: PAW, RCM, AFJ, PJFVB; Critical revision: PAW, RCM, AFJ, PJFVB.

## Abstract

**Aim:** To evaluate the preparedness and adherence of Brazilian long-term care facilities (LTCFs) to the World Health Organization (WHO) infection prevention and control (IPC) guidance and examine the association of LTCF size with adherence to recommendations.

**Methods:** We conducted a cross-sectional study of LTCF managers for 12 consecutive weeks from May 5, 2020. We developed and pre-tested a 46-item questionnaire based on WHO IPC guidance that included multiple-choice and dichotomous questions as well as an open-ended question on the main difficulties encountered by the facility in tackling the pandemic. Using a global adherence score based on the adoption of 20 recommendations, we classified preparedness as (1) **excellent** for LTCFs following ≥14 recommendations, (2) **good** for those following 10-13 recommendations, and (3) **poor** for those following <10 recommendations. LTCF size was established as small, medium, and large according to a 2-step cluster analysis of the number of residents as a continuous variable. We used descriptive statistics and chi-square tests at a 5% significance level.

**Results:** Of 362 facilities included in the study, 308 (85.1%) adhered to 14 or more recommendations; 3 were classified as poorly adherent. Regarding LTCF size, we found a lower adherence to screening visitors for COVID-19 signs and symptoms (p=0.037) and to isolating patients until they have 2 negative laboratory tests (p=0.032) in larger facilities than in medium and small facilities.

**Conclusions:** Preparedness for mitigating COVID-19 in Brazilian LTCFs was considered excellent for most of the proposed recommendations, regardless of LTCF size. Difficulties and problems with infrastructure and/or resident care were much less commonly reported than those related to maintenance of a sufficient stock of materials, workforce management, and financial distress.

## INTRODUCTION

Older people living in long-term care facilities (LTCFs) have been disproportionately affected by the COVID-19 pandemic.^1–4^ Despite considerable variation, mortality rates in LTCFs accounted for more than 70% of all COVID-19 deaths during the first and second waves in some countries.^2,5^ The pandemic has not only overwhelmed health systems worldwide but also placed a spotlight on the weaknesses (or absence) of national datasets for LTCFs.^6^

Prevalence of COVID-19 in the community is a strong predictor of number of cases and deaths in LTCFs.^3,5^ Mortality rates have been markedly increased among the most frail residents, as well as among those living in more crowded facilities.^7^ Moreover, the probability of having COVID-19 cases seems to be higher in large not-for-profit facilities located in metropolitan counties.^3^ Consistent with these findings, Abrams et al., examining the characteristics of LTCFs with documented COVID-19 cases in the United States, found that the cases were related to facility location (i.e., urban) and size (i.e., more than 50 beds).^1^ Conversely, Gorges et al. showed that high nurse aide and total nursing hours contribute to mitigating COVID-19 infection in LTCFs.^3^ High staffing ratios^7^ and early and robust infection prevention and control (IPC) practices^5^ seem to be associated with fewer COVID-19 cases and deaths among older adults living in LTCFs.

Early on in the pandemic, the World Health Organization (WHO), the Centers for Disease Control and Prevention (CDC), and regional organizations published recommendations and policy actions to mitigate the impact of COVID-19 on people who rely on the long-term care sector.^8–11^ To the best of our knowledge, however, COVID-19 preparedness in LTCFs and adherence to recommendations published by international organizations have not been sufficiently investigated.

In a study conducted in the United States with a small sample of LTCFs, the most commonly used IPC guidance was CDC recommendations (88%), followed by state or local health department documents (84%) and WHO guidance (48%).^12^ More than half of LTCF managers (54%) had separate COVID-19 plans, almost all (96%) had policies for screening visitors, and most of them (68%) indicated they had a local referral hospital accepting their patients under investigation for COVID-19. Nearly 83% expected significant staff shortages, while 66% reported access to COVID-19 testing.^12^

Implementation of evidence into clinical practice is neither easy nor fast. Barriers and facilitators need to be addressed promptly and correctly, particularly at the organizational level.^13–15^ It is not clear whether LTCF size may be a barrier to implementing strategies to reduce the spread of COVID-19. In large LTCFs, for example, the greater the number of residents, staff, and visitors, the greater the need for staff training and rotation schedules for potential absenteeism, the larger the number of demands for staff turnover/retention, and the higher the risk of supply shortage.^16,17^

This study aimed to evaluate the preparedness and adherence of Brazilian LTCF managers to WHO IPC guidance and examine the association of LTCF size with adherence to recommendations for COVID-19 mitigation.

## METHODS

Ethical approval was given by the institutional review board of the Botucatu Medical School, São Paulo State University (Unesp) for this cross-sectional study (CAAE 30577520.0.0000.0008, protocol no. 4.012.489), and all participants provided online informed consent. The datasets supporting the findings of this study are available in Wachholz, Patrick; Melo, Ruth Caldeira; Jacinto, Alessandro Ferrari; Villas Boas, Paulo José Fortes, 2021, “COVID-19 in Brazilian Long-Term Care Facilities”, https://doi.org/10.7910/DVN/LXBFQG, Harvard Dataverse, V1, https://dataverse.harvard.edu/dataset.xhtml?persistentId=doi:10.7910/DVN/LXBFQG. For the purposes of this study, LTCF was defined as a facility that provides long-term care and/or rehabilitative, restorative, and end-of-life care to residents in need of assistance with activities of daily living, including a variety of services (social, medical, and personal care) to people who are unable to live independently.

### Design and participants

We conducted this cross-sectional study exclusively through electronic platforms using Google Forms for 12 consecutive weeks from May 5, 2020. Managers of Brazilian LTCFs for older people were the population of interest. We obtained their contact information through listings on domains available on the internet and by active search with health secretariats, epidemiological surveillance agencies, stakeholders, the Brazilian Unified Social Care System, and LTCF support groups, including websites and social media groups. We did not apply restrictions regarding the size, location, or type of facility.

### Procedures and measurements

We developed a 46-item questionnaire based on WHO IPC guidance for LTCFs in the context of COVID-19 published on March 21, 2020, and tested it in a pilot study (Doc S1).^18^ The questionnaire is divided into 9 sections: prevention, physical distancing in the facility, rules for visitors, prospective surveillance for COVID-19 among residents, prospective surveillance among employees, source control, restriction of movement and transport, availability of structure for isolation and provision of personal protective equipment (PPE) and cleaning supplies, and technical support to tackle the pandemic. Of the 46 items, 20 are not part of WHO IPC guidance; however, after consensus among the authors, they were considered both pertinent and essential for understanding the strategies for tackling the pandemic in LTCFs and thereby retained in the questionnaire.

The questionnaire includes multiple-choice and dichotomous questions as well as an open-ended question on the main difficulties encountered by the facility in tackling the pandemic. We created a global score for adherence to WHO IPC guidance based on the adoption of 20 recommendations (Doc S2), as extracted from the original IPC questionnaire. For the purposes of this study, we classified preparedness for COVID-19 as (1) **excellent** for LTCFs following at least 14 of the 20 recommendations (≥70%), (2) **good** for those following 10 to 13 recommendations (50% to 69%), and (3) **poor** for those following less than 10 recommendations (≤49%).

The questionnaire is available as Doc S1. Doc S2 highlights the questions that make up the global adherence score. The estimated time to complete the questionnaire was 30 minutes. Respondents were able to complete the questionnaire more than once, but each LTCF was included only once in the study. That is, where LTCFs had replied more than once, only the most complete and recent questionnaire was included in the analysis.

### Statistical analysis

We analyzed the data in SPSS version 20. When respondents did not report data for one or more variables, we described them as missing data and informed their proportional representation. As there is no standard classification for LTCF size, for the purposes of this study, we performed a 2-step cluster analysis of the number of residents as a continuous variable using the automatic clustering algorithm in SPSS version 20. Three clusters were then established: small (<18 residents), medium (19-75 residents), and large LTCFs (>76 residents) LTCFs. We used descriptive statistics and chi-square tests at a 5% significance level.

We analyzed the responses to the open-ended question using thematic content analysis,^19^ divided into 3 steps: pre-analysis, material exploration, and treatment of results. We first performed a free-floating reading to identify the main difficulties encountered by the LTCFs (Table S1). Subsequently, we explored the material to establish central themes and related subcategories. We then coded the responses in order to calculate each category’s frequency and to extract parts of the related text, thus creating a Venn diagram (Figure 1).

**Figure 1.**
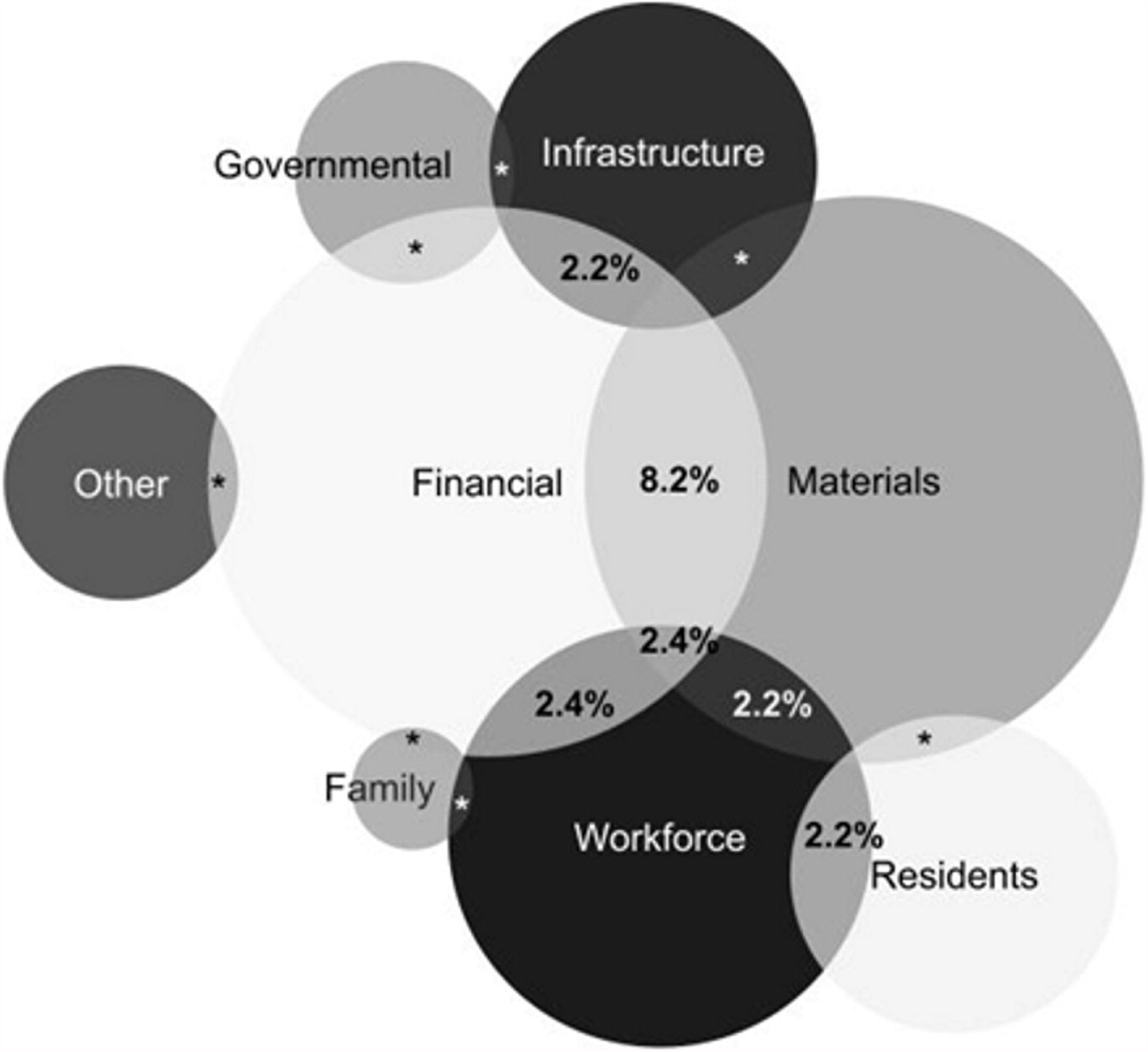
Venn diagram showing the overlap of issues faced by long-term care facilities.

## RESULTS

We received 374 replies during the 12-week investigation period. Five LTCF managers who answered the questionnaire twice had their oldest and less complete responses excluded; one reply from a non-Brazilian LTCF was also excluded. Six managers did not provide data on the number of residents, so their responses were not included. This resulted in 362 LTCFs included in the analysis.

Altogether the 362 LTCFs served a population of 111903 older adults. The Southeast region of Brazil had the largest proportion of included facilities (53.8%), most of which were for-profit LTCFs (39.7%) and not-for-profit LTCFs receiving government/nongovernment funds (37.5%). Table 1 shows the characteristics of the LTCFs included in the study, divided according to facility size.

**Table 1.**
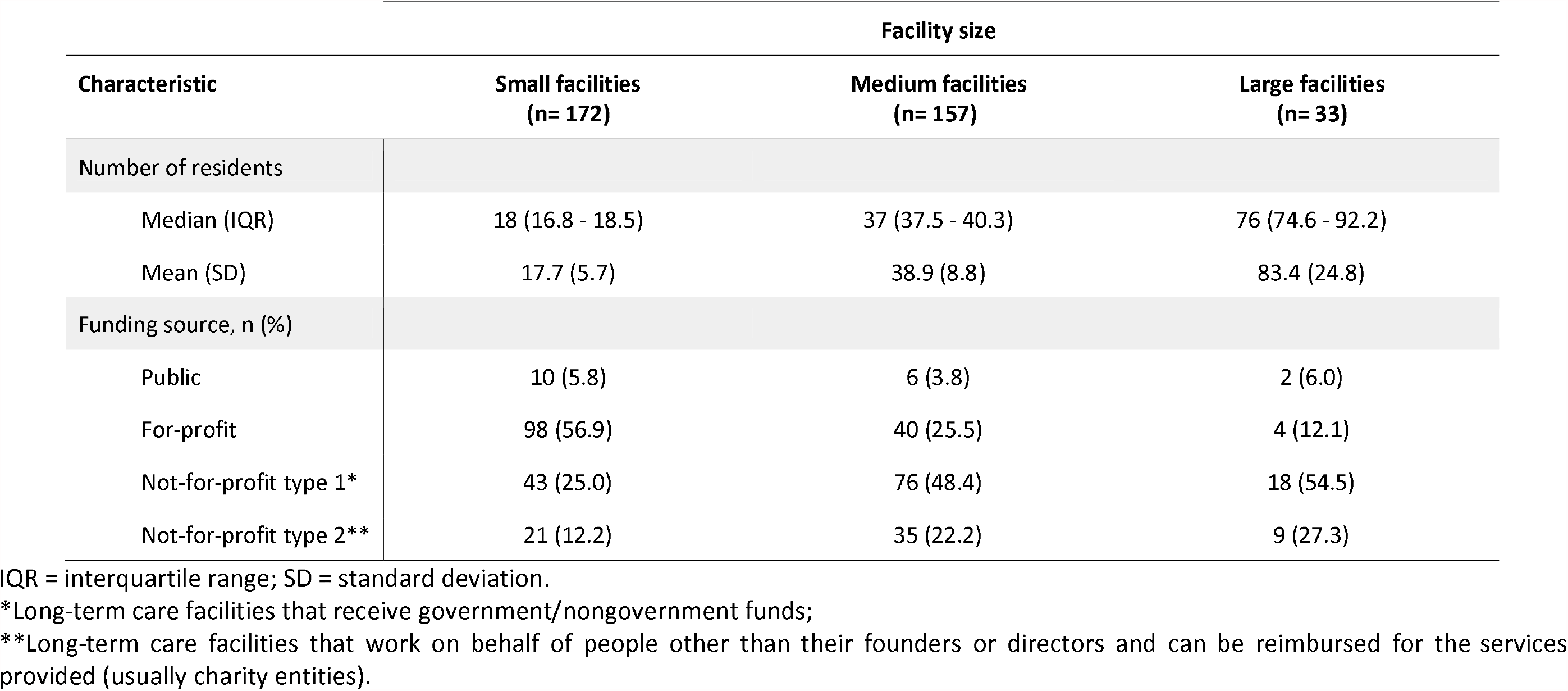
Characteristics of 362 Brazilian long-term care facilities for older people, according to facility size, whose managers answered an online questionnaire to identify their preparedness for the COVID-19 pandemic.

Regarding funding, 166 LTCF managers (45.1%) reported that they had not received any funding or external financial support to prepare themselves to deal with the pandemic, including training, purchasing of PPE, and infrastructure adjustments for respiratory isolation of suspected cases.

Two hundred and thirty-five managers (64.9%; 74 missing data) answered that their facilities already had the necessary infrastructure to accommodate residents with suspected COVID-19, including rooms with a private bathroom and sufficient space for employees and residents to carry out preventive, hygiene, and protection practices.

Access to laboratory tests for influenza and coronavirus was low: 23.5% (n=85) had access to neither, and 17.4% (n=63) had access only to SARS-CoV-2 rapid test kits.

Table 2 summarizes adherence to WHO IPC guidance for LTCFs in the context of COVID-19. Of 362 LTCFs, 308 (85.1%) adhered to 14 or more WHO IPC guidance recommendations; 3 LTCFs were classified as poorly adherent, and 35 (9.7%) had missing data. Regarding LTCF size, we found a lower adherence to screening visitors for COVID-19 signs and symptoms (p=0.037) and to isolating patients until they have 2 negative laboratory tests for COVID-19 (p=0.032) in larger facilities than in medium and small facilities. No significant differences were found in other recommendations between small, medium, and large LTCFs.

**Table 2.**
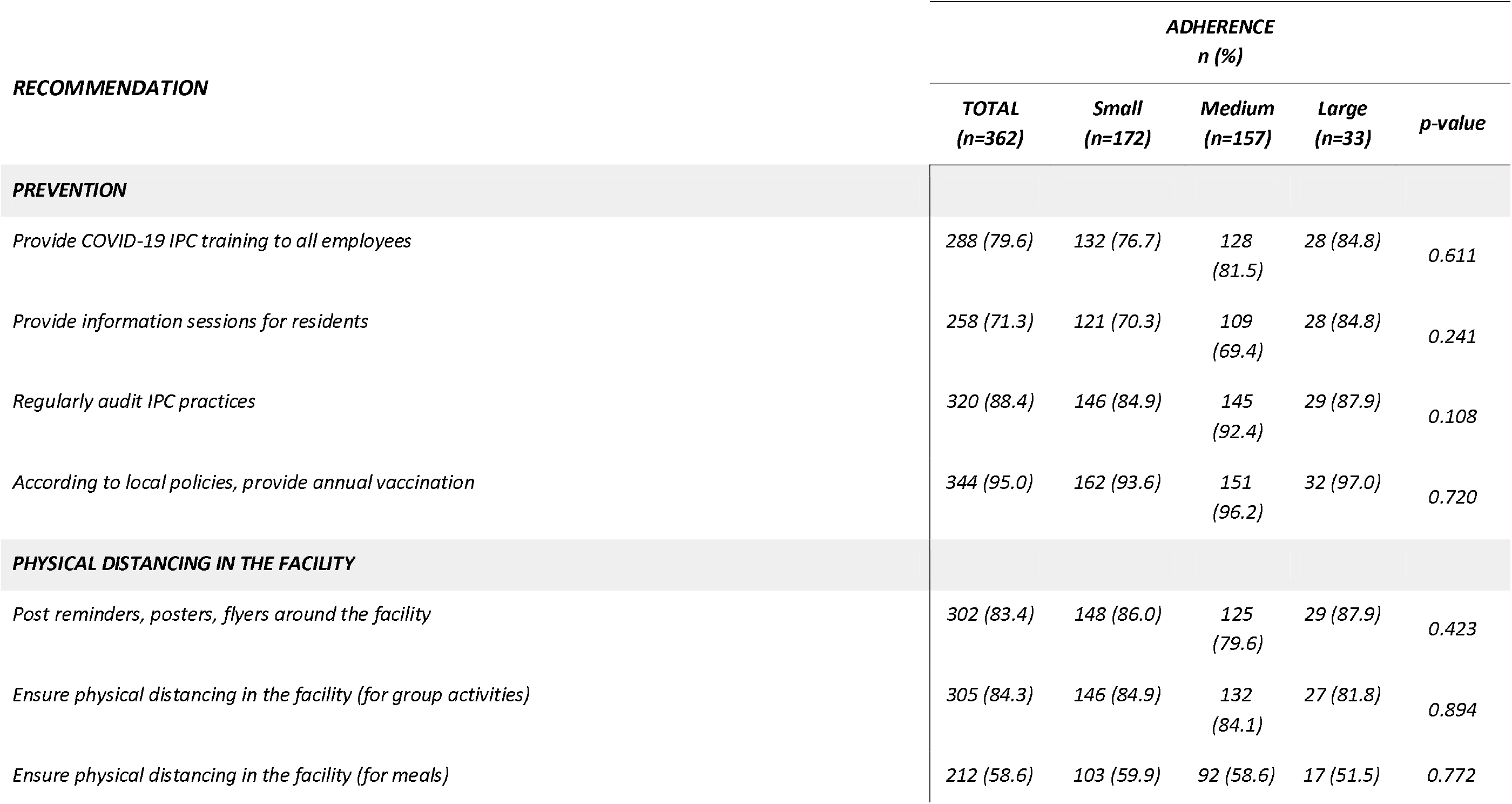
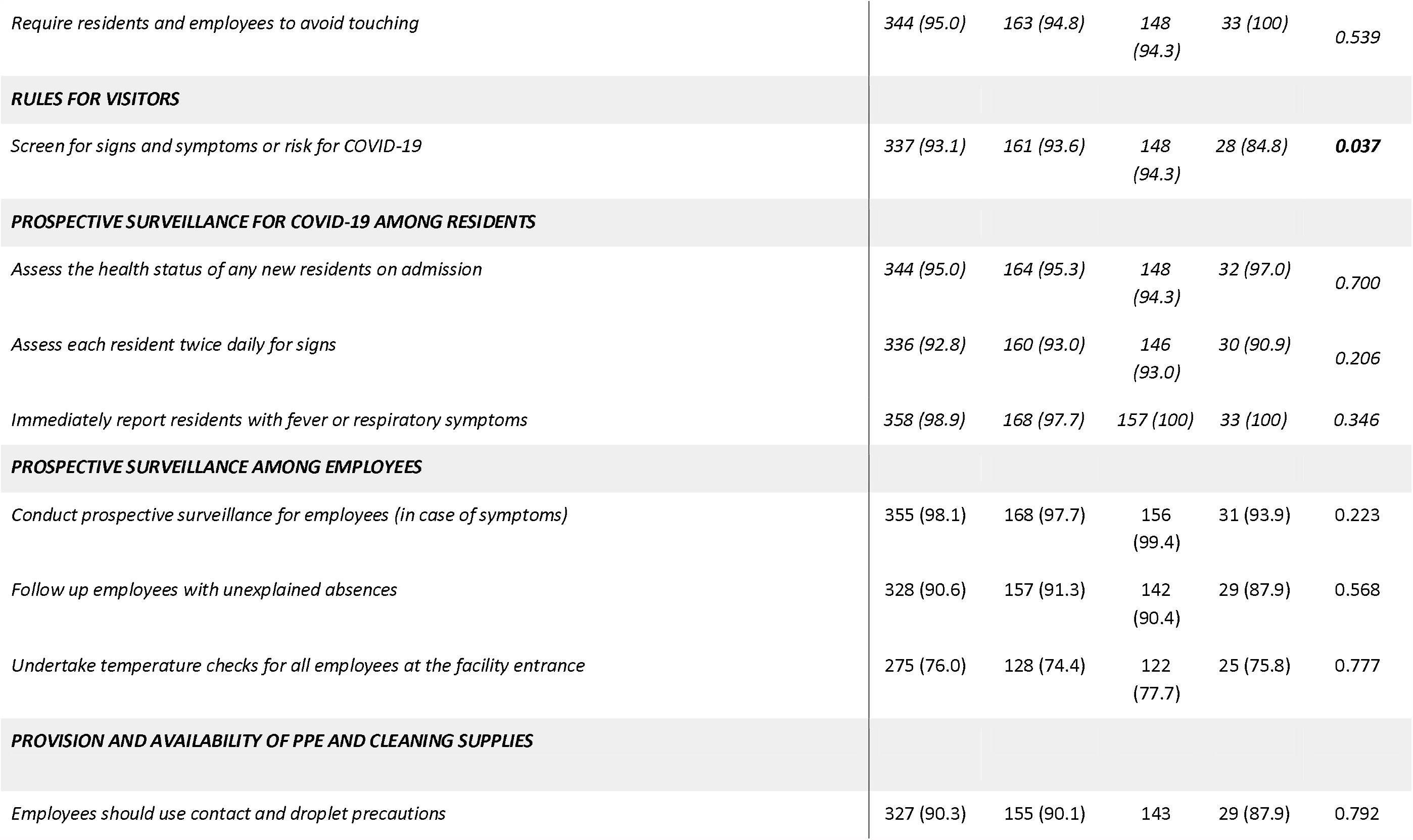
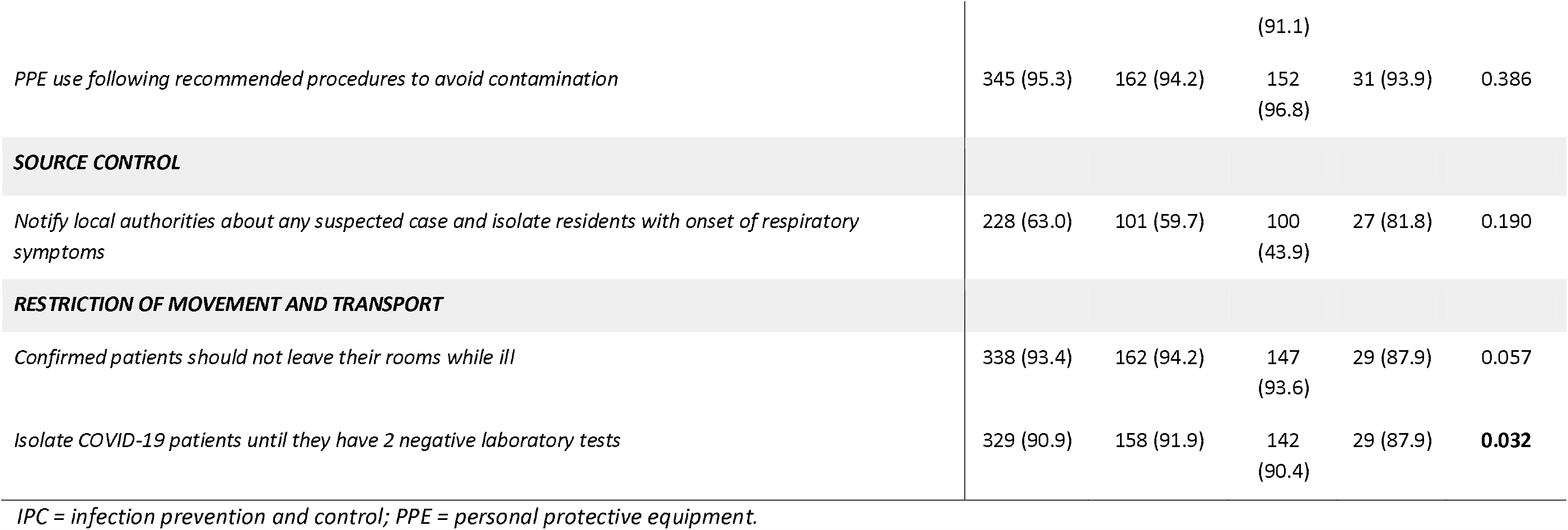
Adherence to the World Health Organization infection prevention and control guidance for long-term care facilities in the context of COVID-19 in 362 Brazilian care homes.

Table 3 describes the issues that could influence preparedness and adherence to additional technical support recommendations to deal with the COVID-19 pandemic in LTCFs, involving questions not included in the global adherence score. The most common issue was lack of external support (74.3% did not receive support to plan and execute training and contingency plans), followed by difficulty in purchasing PPE for residents and employees (47.0%) and managing deaths within the facility (46.1%). The creation of a multidisciplinary planning committee specifically to deal with COVID-19 issues was less common in small LTCFs (49.4%) than in medium (65.0%) and large (75.8%) facilities (p=0.034).

**Table 3.** Preparedness and adherence to technical support recommendations to tackle the COVID-19 pandemic in 362 Brazilian long-term care facilities.

| RECOMMENDATION | ADHERENCE<br>n (%) |  |  |  | p-value |
| --- | --- | --- | --- | --- | --- |
|  | TOTAL<br>(n=362) | Small<br>(n=172) | Medium<br>(n=157) | Large<br>(n=33) |  |
| **CONTINGENCY PLAN** |  |  |  |  |  |
| *Managers have discussed, analyzed, or considered developing a contingency plan to identify the minimum number of employees needed to operate safely.* | 276 (72.7) | 128 (74.4) | 124 (79.0) | 24 (72.7) | 0.838 |
| *Managers have effectively implemented a contingency plan to identify the minimum number of employees needed to operate safely and on how to hire or recruit them.* | 241 (66.6) | 110 (64.0) | 109 (69.4) | 22 (66.7) | 0.304 |
| *A multidisciplinary planning committee has been created specifically to decide and discuss the planning of actions to prevent and combat COVID-19.* | 212 (58.6) | 85 (49.4) | 102 (65.0) | 25 (75.8) | **0.034** |
| **PERSONAL PROTECTIVE EQUIPMENT (PPE)** |  |  |  |  |  |
| *Residents and employees have access to comfortable and sufficient PPE.* | 323 (89.2) | 156 (90.7) | 137 (87.3) | 30 (90.9) | 0.716 |
| *The facility is having difficulty in purchasing or stocking cleaning supplies and PPE for residents and employees.* | 192 (53.0) | 89 (51.7) | 89 (56.7) | 14 (42.2) | 0.499 |

| **MANAGEMENT OF SUSPECTED/CONFIRMED CASES AND DEATHS** |  |  |  |  |  |
| --- | --- | --- | --- | --- | --- |
| *The facility has established care flows with local health authorities or referral units to transfer suspected cases.* | 228 (63.0) | 101 (58.7) | 100 (63.7) | 27 (81.8) | 0.199 |
| *Managers have developed a contingency plan to deal with deaths within the facility.* | 195 (53.9) | 80 (46.5) | 95 (60.5) | 20 (60.6) | 0.249 |
| **EXTERNAL SUPPORT** |  |  |  |  |  |
| *The facility has received funding or external support to plan and execute the training and contingency plans needed to deal with the COVID-19 pandemic.* | 93 (25.7) | 33 (19.2) | 48 (30.6) | 12 (36.4) | 0.169 |

The analysis of managers’ responses to the open-ended question showed that most LTCFs (98%) were having difficulty in tackling the COVID-19 pandemic. According to the managers, the most affected areas were those related to keeping a sufficient stock of materials (42%) (e.g., PPE, hygiene and cleaning supplies, and COVID-19 tests), financial distress (39%) (e.g., extra expenses with materials, additional costs with staff, and fundraising issues), and managing the workforce (24%) (e.g., absence and replacement, awareness and protocol compliance, qualification and training, and emotional distress). In 14% of LTCFs, managers reported problems related to infrastructure (e.g., adjustments for isolation of suspected/confirmed cases and enough space to ensure physical distancing) and to residents (e.g., awareness and protocol compliance, emotional distress, compliance with social distancing, management of dementia, restriction of group activities and visitors, and lack of family contact). Detailed results are available as Supplementary file 3.

Figure 1 illustrates the overlap of issues faced by LTCFs, according to their managers. The main overlap was detected between financial and materials issues (8.2%), followed by financial vs workforce (2.4%) and financial vs materials vs workforce (2.4%). A 2.2% of overlap was also observed for financial vs infrastructure, materials vs workforce, and workforce vs residents. The remaining overlaps of issues faced by LTCFs were present in less than 2.0% of responses.

## DISCUSSION

In this cross-sectional study, most Brazilian LTCFs (85.1%) reported high adherence to WHO IPC guidance to mitigate COVID-19, albeit 98% reported having difficulties with shortage of supplies, PPE, and materials, workforce management, and financial problems.

Although previous evidence clearly suggests that LTCF size is associated with an increased risk of outbreaks and deaths, and possibly with difficulty adhering to IPC measures,^1,3^ we found a high overall adherence rate regardless of LTCF size. The global adherence score was slightly lower in larger LTCFs for ‘screening external visitors’ and ‘isolating infected residents’. Adherence to the recommendation of establishing multidisciplinary committees to combat COVID-19 decreased with decreasing LTCF size.

Although the size of facilities was not assessed in a recent systematic review on the epidemiology and clinical features of COVID-19 outbreaks in aged care facilities,^20^ larger facilities had been previously found to be correlated with the spread of infection.^21,22^ However, increasing testing capacity and updating surveillance protocols accordingly could facilitate earlier detection of emerging outbreaks and help these facilities manage the supply of care workers and quality of nursing homes, especially their response to infectious diseases.^21^

Many LTCFs have perceived a considerably high workload during the COVID-19 pandemic. Tasks and activities that require direct involvement of the workforce, such as caring for infected residents (particularly those with functional impairment and cognitive decline) and screening external visitors, would be expected to have the most significant impact on facilities with the highest number of residents. This impact, however, was not sufficient to reduce adherence to IPC guidance significantly.

In the United States, COVID-19 mortality was found to be higher in significantly more crowded facilities (9.7%) than in less crowded ones (4.5%), regardless of facility size.^23^ The size of LTCFs was strongly associated with COVID-19 outbreaks (odds ratio per 20-bed increase 3.35, 95% confidence interval 1.99–5.63).^24^ Meanwhile, in the UK, the likelihood of spread of COVID-19 was higher for larger LTCFs (>20 beds) and when workers and facilities did not adhere to IPC measures to mitigate infection risk.^25^ Preparedness and adherence to recommendations by LTCFs, however, have been poorly described in low- and middle income countries (LMICs).

Prevention is a critical component of infection control, particularly during emerging outbreaks. The long-term care sector in LMICs remains mostly underdeveloped, and specific strategies must be considered and suited to promote and successfully implement IPC protocols and guidelines. In a study of high-risk populations in a high-income country, IPC implementation significantly differed between higher- and lower-prevalence groups in the social distancing and PPE categories.^23^

Some factors can potentially influence adherence to IPC guidance by acting as facilitators or barriers to its implementation. The community prevalence of COVID-19, the availability of screening tests, and the rate of infection among workers (including staff turnover and retention) may influence adherence to some of the IPC recommendations. Other factors that may affect adherence include the facility’s occupancy rate, inadequate staff IPC measures to minimize staff-to-staff transmission, delayed recognition of cases in residents because of a low index of suspicion, and residents at risk for severe morbidity and death sharing a location.^25^

Despite the lack of official data, COVID-19 mortality in Brazilian LTCFs is lower than in other countries.^6^ Although the early suspension of visits to LTCFs has been hypothesized to influence these rates, there is no robust evidence to support this statement at the moment. The adoption of good practices to mitigate the impact of COVID-19, such as training targeting the workforce, use of PPE, and adherence to the IPC guidance, may have influenced the results; however, these results still need confirmation by longitudinal studies. Previous studies involving 23⍰896 Brazilian respondents (mean age, 47.4⍰years) have identified a satisfactory level of adherence to national COVID-19 prevention guidelines. Younger people, males, persons living in rural areas/villages or in densely populated or low-income neighborhoods, students, and workers often engage in less preventive behavior.^26^

This study has limitations inherent in the cross-sectional design adopted and the potential for recall bias. Likewise, potential selection bias may have favored LTCFs with access to the internet and those with more complete teams, including workers dedicated to administrative tasks. The instrument used for data collection was tested in a pilot study,^18^ but it is not a validated instrument. Also, there are no established cutoff points to classify facilities by size, which may have influenced the findings. Nevertheless, all definitions adopted for the purpose of this study were defined a priori, and a large and nationally representative number of LTCFs was included.

COVID-19 has been a pandemic, or a syndemic,^27^ of inequalities. Countries with successful responses, for instance, developed partnerships on multiple levels across government sectors, had timely triage and referral of suspected COVID-19 cases, and provided designated isolation facilities.^28^ The drama observed at the beginning of the pandemic, when COVID-19 deaths among LTCF residents were brutally high in some countries, was partly due to the lack of official guidelines and regulations for natural disasters and pandemics worldwide.^29^ Researchers found that only 57% of 182 countries could perform crucial activities to prevent, detect, and respond to an outbreak in the context of COVID-19.^30^ As previously stated, context matters.^27^

Coronavirus outbreaks will probably continue to occur worldwide for the following years, similar to other respiratory viruses.^23,27^ The effects of vaccination and immunity rates remain largely unknown, and they may be lower in LTCF residents due to immunosenescence.^28^

In conclusion, based on adherence to WHO IPC guidance, preparedness for mitigating COVID-19 in Brazilian LTCFs in this study was considered excellent for most of the proposed recommendations, regardless of LTCF size. Difficulties and problems with infrastructure and/or resident care were much less commonly reported than those related to maintenance of a sufficient stock of materials, workforce management, and financial distress.

## Supporting information

Supplemental DocS2

Supplemental DocS1

## Data Availability

The datasets supporting the findings of this study are available in Wachholz, Patrick; Melo, Ruth Caldeira; Jacinto, Alessandro Ferrari; Villas Boas, Paulo Jose Fortes, 2021, "COVID-19 in Brazilian Long-Term Care Facilities", https://doi.org/10.7910/DVN/LXBFQG, Harvard Dataverse, V1, UNF:6:ZmzY+hPU0gmqmos34oPxJA== [fileUNF].

https://dataverse.harvard.edu/dataset.xhtml?persistentId=doi:10.7910/DVN/LXBFQG

## Acknowledgments

We thank researcher Helena Akemi Wada Watanabe, the ‘Frente Nacional de Fortalecimento às ILPIs,’ and the group of professionals and researchers from the ilpi.me website for their support in carrying out this study.

## Supporting information

Doc S1 - COMPLETE QUESTIONNAIRE

Doc S2 - QUESTIONS INCLUDED IN THE GLOBAL SCORE FOR ADHERENCE TO WHO IPC GUIDANCE

Table S1 - “Free-floating reading” coding on the main difficulties encountered by LTCFs in coping with the COVID-19 pandemic.

