## Supplemental DocS2 for "Impact of long-term care facility size on preparedness and adherence to infection prevention and control guidance for the mitigation of COVID-19"

**DOC S2 – QUESTIONS INCLUDED IN THE GLOBAL SCORE FOR ADHERENCE TO WHO IPC GUIDANCE**

1.1) At your facility, training based on the infection prevention and control guidance for the novel coronavirus has been provided to all employees, including at least the following topics: general information about the coronavirus; hand hygiene and behavior to be adopted in the presence of respiratory symptoms (coughing, sneezing, etc.); standard contact precautions; and virus transmission-based precautions. Please select the option that best represents the type of training provided at your facility:

•	Training already carried out

YES

NO

1.2) Information meetings have been held with residents who are able to minimally understand about COVID-19 to inform them about the virus, symptoms, and diseases caused by the virus and how to protect themselves from infection.

•	Completed

•	Not completed

1.3) Infection prevention and control practices (e.g., hand hygiene compliance) and review of hygiene and cleaning standards have been regularly encouraged and checked, particularly with employees (and residents), and feedback has been provided to employees.

•	Yes

•	No

1.4) Have annual influenza vaccination (seasonal flu and H1N1) and antipneumococcal vaccines [according to local health policies] been made available and recommended for employees and residents?

\# WHO REMINDER: Influenza and pneumococcal infections (preventable by vaccination) are often associated with respiratory mortality in older people, regardless of COVID-19.

•	Completed

YES

NO

3.1) All visitors are minimally questioned and/or, if feasible, physically examined for signs and symptoms of acute respiratory infection (fever, cough, shortness of breath, diarrhea, etc.) or significant risk for COVID-19 (contact with someone with these symptoms in the last 7-14 days). No one presenting with these characteristics has been allowed to enter the premises.

•	Yes

•	No

4.1) The health status of new residents has been assessed prior to admission to the facility to determine whether the resident has signs or symptoms of respiratory disease, OR no new admissions have been accepted in the facility during the pandemic.

•	Yes

•	No

4.2) Each resident has been assessed periodically (ideally twice a day) for the development of fever, cough, or difficulty breathing.

•	Yes

•	No

4.3) Cases of residents with fever or respiratory symptoms have been promptly reported to the staff responsible for clinical follow-up (leader / head of the nursing staff, physician, or other health professionals) and to the local health department responsible for the follow-up and monitoring of suspected COVID-19 cases in the municipality or region.

•	Yes

•	No

5.2) Employees with unexcused absences have been followed up, and their health status has been actively monitored (by phone, for example), as well as that of their families, in order to find out if someone may be developing infection symptoms, especially if there is information of contact with suspected or confirmed COVID-19 cases.

•	Yes

•	No

5.3) Employees' body temperature has been checked before entering the facility, and visibly sick employees have been referred for evaluation by their local health care provider.

•	Yes

•	No

6.2) When providing routine care to a resident with suspected (or confirmed) COVID-19, all contact and droplet precautions have been implemented and personal protective equipment has been provided to the employees.

•	Yes

•	No

6.3) Employees have been instructed and/or are constantly reminded to appropriately remove their personal protective equipment upon leaving the environment where they have been taking care of a resident with suspected (or confirmed) COVID-19, to dispose of the equipment immediately in the appropriate waste bins, and to perform hand hygiene. That is, they discard gloves, cap, gown, mask, and any other clothing that may have been in contact with secretions and then perform hand hygiene practices.

•	Yes

•	No

7.1) Employees, residents, and family members have been informed that residents with suspected and/or confirmed COVID-19 CANNOT leave their rooms while they are ill (therefore, they must have access to a private toilet).

\# WHO REMINDER: The virus may be transmitted through feces in addition to respiratory secretions.

•	Yes

•	No

7.2) Residents with suspected COVID-19 are kept in isolation until they have 2 negative laboratory tests for COVID-19 performed at least 24 h apart (when available from the local health system) OR until all symptoms have resolved (minimum of 2 weeks).

•	Yes

•	No

8.6) If there is a need to transfer residents with suspected COVID-19, the facility has already discussed or established contingency protocols and care flows together with the local health authorities, primary health care units, referral hospital, or mobile urgency and emergency services.

•	Yes

•	No
