## Supplemental DocS1 for "Impact of long-term care facility size on preparedness and adherence to infection prevention and control guidance for the mitigation of COVID-19"

DOC S1 – COMPLETE QUESTIONNAIRE

STEP 1 - ANONYMOUS DATA

The data collected in this step will be coded and excluded from the final records to prevent the responses from being identified.

As described in the Informed Consent Form, your responses are confidential and will be used exclusively for the purposes previously described for this investigation.

If you wish, you can receive your responses via email. If so, activate the option to send a reply when you finish completing the questionnaire.

1) Please describe how many residents live in the facility: _____

2) Please select the type of facility:

•	Public

•	Private

•	Philanthropic that charges tuition or fees

•	Nonprofit philanthropic (receives government or nongovernment funds)

•	Other

3) Please describe the location of the facility (by location, we mean CITY, STATE / PROVINCE, COUNTRY). Example: São Paulo, SP, Brazil: ___________________________________

4) Please inform the MAIN source of information the facility has employed to adapt and prepare for the COVID-19 pandemic?

•	None

•	Government

•	Nongovernmental institutions

•	Scientific associations

•	Other sources

If you selected "Other sources" in the previous item, please specify: ____________________

STEP 2 - WHAT HAS BEEN DONE

On March 21, 2020, the World Health Organization (WHO) published international guideline recommendations for tackling the COVID-19 pandemic in long-term care facilities for older people. The following questions are based on this guidance.

These questions will NOT be used to label facilities nor to create any ranking among them. They are not intended to be sufficient to assess the whole subject related to COVID-19.

They can, however, be an exciting and valuable guide for managers who wish to use them to check whether the measures they have been adopting to protect their employees and residents are in line with the international recommendations made by WHO.

1) PREVENTION

1.1) At your facility, training based on the infection prevention and control guidance for the novel coronavirus has been provided to all employees, including at least the following topics: general information about the coronavirus; hand hygiene and behavior to be adopted in the presence of respiratory symptoms (coughing, sneezing, etc.); standard contact precautions; and virus transmission-based precautions. Please select the option that best represents the type of training provided at your facility:

•	Pieces of training already carried out (COMPLETED)

•	Training not carried out (IN PROGRESS / NOT STARTED / NO TRAINING / THESE TOPICS WERE NOT INCLUDED / NA)

1.2) Information meetings have been held with residents who are able to minimally understand about COVID-19 to inform them about the virus, symptoms, and diseases caused by the virus and how to protect themselves from infection.

•	Completed

•	Not completed

1.3) Infection prevention and control practices (e.g., hand hygiene compliance) and review of hygiene and cleaning standards have been regularly encouraged and checked, particularly with employees (and residents), and feedback has been provided to employees.

•	Yes

•	No

1.4) Have annual influenza vaccination (seasonal flu and H1N1) and antipneumococcal vaccines [according to local health policies] been made available and recommended for employees and residents?

\# WHO REMINDER: Influenza and pneumococcal infections (preventable by vaccination) are often associated with respiratory mortality in older people, regardless of COVID-19.

•	Completed (all residents and employees were vaccinated)

•	Not completed (in progress / not started / NA)

2) PHYSICAL DISTANCING IN THE FACILITY

2.1) In group activities with residents, the recommended physical distance between residents (minimum of 1 meter; 1.5 meters if possible) has been ensured. If not feasible, group activities have been canceled.

•	Yes

•	No

•	Partially

•	Do not know

2.2) Meals have been staggered (that is, residents have been divided into groups at different mealtimes) to ensure that the minimum physical distance between residents is maintained. If not feasible, dining halls have been closed and meals started to be served individually in the residents' accommodations.

•	Yes

•	No

•	Partially

•	Do not know

2.3) Residents and employees have been instructed to avoid physical contact, and guidance on the importance of maintaining a minimum 1-meter distance has been constantly reinforced for both.

•	Yes

•	No

•	Partially

•	Do not know

2.4) Signs have been posted around the facility to remind residents and employees of the recommendations for physical distancing and infection prevention (covering the mouth when coughing, hand washing, avoiding touching eyes/mouth/nose with hands, etc.)

•	Yes

•	No

•	Do not know

3) RULES FOR VISITORS

3.1) All visitors are minimally questioned and/or, if feasible, physically examined for signs and symptoms of acute respiratory infection (fever, cough, shortness of breath, diarrhea, etc.) or significant risk for COVID-19 (contact with someone with these symptoms in the last 7-14 days). No one presenting with these characteristics has been allowed to enter the premises.

•	Yes

•	No

4) PROSPECTIVE SURVEILLANCE FOR COVID-19 AMONG RESIDENTS

4.1) The health status of new residents has been assessed prior to admission to the facility to determine whether the resident has signs or symptoms of respiratory disease, OR no new admissions have been accepted in the facility during the pandemic.

•	Yes

•	No

4.2) Each resident has been assessed periodically (ideally twice a day) for the development of fever, cough, or difficulty breathing.

•	Yes

•	No

4.3) Cases of residents with fever or respiratory symptoms have been promptly reported to the staff responsible for clinical follow-up (leader / head of the nursing staff, physician, or other health professionals) and to the local health department responsible for the follow-up and monitoring of suspected COVID-19 cases in the municipality or region.

•	Yes

•	No

5) PROSPECTIVE SURVEILLANCE AMONG EMPLOYEES

5.1) Employees have been actively consulted and encouraged to report if they have fever or respiratory symptoms and are advised not to go to work if they have these symptoms, as a non-punitive, flexible sick leave policy.

•	Yes

•	No

•	Partially

•	Do not know

5.2) Employees with unexcused absences have been followed up, and their health status has been actively monitored (by phone, for example), as well as that of their families, in order to find out if someone may be developing infection symptoms, especially if there is information of contact with suspected or confirmed COVID-19 cases.

•	Yes

•	No

5.3) Employees' body temperature has been checked before entering the facility, and visibly sick employees have been referred for evaluation by their local health care provider.

•	Yes

•	No

5.4) The development of a contingency plan has been discussed, analyzed, or considered in order to identify the minimum number of employees needed for the facility to operate safely (in the case of employees leaving) and to provide guidelines on how to hire or recruit extra staff if necessary.

•	Yes

•	No

•	Partially

•	Do not know

5.5) A contingency plan has been implemented to identify the minimum number of employees needed for the facility to operate safely (in the case of employees leaving) and to hire or recruit extra staff if necessary.

•	Yes

•	No

•	Partially

•	Do not know

6) SOURCE CONTROL

6.1) If your facility has identified (or might identify a suspected COVID-19 case), which local authorities have been (or should be) notified of these cases?

•	Epidemiological surveillance agency

•	Public prosecutor’s office

•	Local operational support center for coping with COVID-19

•	Do not know

•	Other: _______________________________

6.2) When providing routine care to a resident with suspected (or confirmed) COVID-19, all contact and droplet precautions have been implemented and personal protective equipment has been provided to the employees.

•	Yes

•	No

6.3) Employees have been instructed and/or are constantly reminded to appropriately remove their personal protective equipment upon leaving the environment where they have been taking care of a resident with suspected (or confirmed) COVID-19, to dispose of the equipment immediately in the appropriate waste bins, and to perform hand hygiene. That is, they discard gloves, cap, gown, mask, and any other clothing that may have been in contact with secretions and then perform hand hygiene practices.

•	Yes

•	No

7) RESTRICTION OF MOVEMENT AND TRANSPORT

7.1) Employees, residents, and family members have been informed that residents with suspected and/or confirmed COVID-19 CANNOT leave their rooms while they are ill (therefore, they must have access to a private toilet).

\# WHO REMINDER: The virus may be transmitted through feces in addition to respiratory secretions.

•	Yes

•	No

7.2) Residents with suspected COVID-19 are kept in isolation until they have 2 negative laboratory tests for COVID-19 performed at least 24 h apart (when available from the local health system) OR until all symptoms have resolved (minimum of 2 weeks).

•	Yes

•	No

7.3) Please describe what restriction of free movement has been implemented within the facility (if you have not yet had a suspected case in the facility) and for residents with memory problems (such as Alzheimer’s disease).

Your answer: _____________________________

8) AVAILABILITY OF STRUCTURE FOR ISOLATION AND PROVISION OF INDIVIDUAL PROTECTIVE EQUIPMENT AND CLEANING SUPPLIES

8.1) Do employees (and residents if necessary) have access to personal protective equipment at your facility?

•	Yes

•	No

•	Partially

•	Do not know

If YES or PARTIALLY, which item(s)?

•	Disposable masks

•	Disposable gloves

•	Alcohol-based hand rubs

•	Sink with soap and clean water

•	Face shield

•	Disposable gown

•	Waterproof boots

•	Other

If you selected “Other”, please specify: _________________________________________

8.2) TODAY, considering the lowest stock of any of the personal protective equipment items mentioned above, you believe that this stock will last for:

•	We are already running short of items

•	A few days

•	No more than 1 week

•	2 to 4 weeks

•	5 to 6 weeks

•	More than 6 weeks

8.3) The facility has found it difficult to purchase or keep the stocks of cleaning supplies and personal protective equipment during the pandemic.

•	Yes

•	No

•	Partially

•	Do not know

8.4) Has your facility had access to laboratory tests to confirm coronavirus infection in suspected cases? Or to tests for influenza (seasonal flu / H1N1)?

•	Yes, for both

•	No, for none of them

•	Influenza only

•	Coronavirus only

•	Do not know

8.5) If necessary, TODAY the facility has available space and infrastructure for respiratory isolation of residents with suspected COVID-19 (that is, it has a room, separated from the accommodations of other residents, with a private bathroom, where older people with symptoms can be assisted and employees can practice the precautions for COVID-19, such as putting on and removing personal protective equipment and performing hand hygiene).

•	Yes

•	No

•	Partially

•	Do not know

8.6) If there is a need to transfer residents with suspected COVID-19, the facility has already discussed or established contingency protocols and care flows together with the local health authorities, primary health care units, referral hospital, or mobile urgency and emergency services.

•	Yes

•	No

8.7) A contingency plan has been developed or at least discussed to deal with eventual deaths within the facility (for example, a specific area has been defined to keep dead bodies and any materials/furniture that may be contaminated, thus reducing the likelihood of contamination of other residents, employees, and visitors).

•	Yes

•	No

•	Partially

•	Do not know

9) TECHNICAL SUPPORT FROM HEALTH AUTHORITIES OR HEALTH SECTOR TO TACKLE THE PANDEMIC

9.1) Has your facility received external support (including financial assistance) or any support for training and preparation for coping with the pandemic?

•	Yes

•	No

•	Partially

•	Do not know

9.2) Has a multidisciplinary planning committee or group been created at your facility specifically to decide and discuss the planning of actions to prevent and combat COVID-19?

•	Yes

•	No

•	Do not know

9.3) How long (in days) has it taken your facility to prepare for tackling the pandemic?

_______________________________________

9.4) IN THE FUTURE, how long (in days) do you believe it would take your facility to adequately prepare for tackling a similar situation, considering what today you identify and perceive as the facility's greatest difficulties?

________________________________________

9.5) What are the MAJOR DIFFICULTIES that your facility encounters today to tackle the pandemic?

________________________________________

9.6) Has your facility prepared a written strategic plan for coping with COVID-19?

•	Completed

•	In progress

•	Not started

•	Do not know

9.7) Does your facility (or the COVID-19 strategic coping group) constantly monitor the publication of new public recommendations on the condition and consider adapting or modifying the action plan based on these recommendations?

•	Yes

•	No

•	Partially

•	Do not know

9.8) Do you know how many residents of your facility died (from any cause) in 2019?

______________________________________

9.9) Of these, do you know how many died of PNEUMONIA only?

______________________________________

9.10) Do you know how many residents of your facility were hospitalized (for any reason) in 2019?

_______________________________________

9.11) Of these, do you know how many were hospitalized for PNEUMONIA?

_______________________________________

9.12) And in 2018, do you know how many residents of your facility died (from any cause)?

_____________________________________________________________________________

9.13) And in 2018, do you know how many residents of your facility were hospitalized (for any reason)?

____________________________________________________________________________
