## Supplemental Table S1 for "Impact of long-term care facility size on preparedness and adherence to infection prevention and control guidance for the mitigation of COVID-19"

Table S1 - "Free-floating reading" used in the analysis of the open-ended question on the main difficulties encountered by LTCFs in coping with the COVID-19 pandemic. Coding and subcoding using Altas.ti

| **Category** | **Subcategory** | **Example** |
| --- | --- | --- |
| Infrastructure | General | "We lack physical structure and financial resources, and the number of employees is insufficient." |
|  | Respiratory isolation | "One of the greatest difficulties is space. All rooms are small, with the minimum distance required between the beds. All rooms are shared, and there is no room for respiratory isolation."… "We lack isolation rooms for suspected or confirmed COVID-19 cases."… "Where to isolate this older resident?" |
| Financial | Extra cost (material) | "The greatest difficulty we are facing is the lack of money to purchase PPE, as it has a high cost that is outside our budget, which makes it very difficult." |
|  | Extra cost (staff) | "Shortage of funding to hire extra staff to improve the monitoring of residents in isolation in the facility." |
|  | Fundraising | "Having the financial resources to maintain the facility given that we need donations for our operation, and in this pandemic they have been reduced considerably." |
|  | New admissions | "The reduction in financial resources and the increase in expenses, since we chose not to admit new residents for the safety of those who are already here." |
|  | Default | "Financial support, because, with the isolation, families have been unable to pay the full monthly fee." |
|  | General | "It is difficult to equalize our financial situation to meet the health requirements because every day there is new information about COVID-19, and we need make adjustments again." |
| Materials | PPE | "Difficulty having access to PPE, because, in addition to the exorbitant price charged for PPE, it is difficult to find and purchase them." |
|  | Cleaning and hygiene | "Shortage of cleaning supplies (alcohol-based hand rubs and other disposable items)." |
|  | COVID-19 tests | "The lack of tests for employees who have flu-like symptoms to ensure that they are not carriers and transmitters." |
|  | General | "Lack of materials to replenish stocks." |
| Workforce | Absence and turnover | "Replacement of staff who need to leave for a while due to the pandemic." |
|  | Awareness and protocol compliance | "Employee awareness" (Ix), "Making employees comply with the protocols established in the facility (1y)." |
|  | Insufficient | "Understaffing." |
|  | Management | "Some staff who work in hospitals had their contracts suspended until the end of the pandemic. This made it difficult to relocate employees." |
|  | Mobility | "The movement of employees, who go to their homes, live with other people, use public transport, ..." |
|  | Qualification and training | "... technical unpreparedness of professionals for coping with the pandemic." |
|  | Emotional distress | "Emotional control of employees and residents." |
| Residents | Awareness and protocol compliance | "The awareness of other residents to practice social distancing when necessary." |
|  | Emotional distress | "... managing the emotional damage resulting from the absence of visitors and the prohibition of leaving the facility. |
|  | Physical distancing | "... it is hard to explain the need to maintain physical distance between residents..." |
|  | Management of dementia | "Social isolation for older residents with functional decline and dementia." |
|  | New admissions, hospitalization, and readmission | "...admission of new residents ... "," ... Referral of serious cases with the possibility of cure; criteria to return to the facility." |
|  | Social, cultural, and physical activities | "Prohibition of visiting family members and engaging in complementary activities for residents (music, crafts, physical activity, etc.)." |
|  | Restriction of visitors and lack of family contact | "The isolation of the residents family. Not being able to see their families is very, very painful for them." |
| Family | Awareness and protocol compliance | "Family members' lack of understanding of the restriction of visitors for residents." |
|  | Emotional distress | "Anxiety of family members, lucid residents, and staff." |
| Governmental | Support | "Municipal, state, federal support. The facility faces all difficulties alone, the ones that have already arisen and so many others that are yet to come." |
| Other | - | "Going out with the resident for routine appointments, we have to be very careful,…" "Going out to buy food and diapers." "The lack of health training for managers in general." |
